# Cognitive trajectories and incident atrial fibrillation in the ASPREE cohort

**DOI:** 10.1101/2025.08.01.25332579

**Authors:** Amy Brodtmann, Nicholas Cheng, Emilio Werden, Johannes Neumann, Catherine Robb, Sharyn M Fitzgerald, Rory Wolfe, Andrew M Tonkin, Suzanne G. Orchard, Joanne Ryan, Robyn L Woods, Caroline Gao, John J McNeil

**Affiliations:** Cognitive Health Initiative, Department of Neuroscience, School of Translational Medicine, Monash University, Melbourne, Australia; Turner Institute for Brain and Mental Health, Monash University, Clayton, Australia; Dementia Mission, The Florey Institute of Neuroscience and Mental Health, Melbourne, Australia; Department of Cardiology, University Heart & Vascular Center Hamburg, university Medical Center Hamburg-Eppendorf, Germany; School of Public Health and Preventive Medicine, Monash University, Melbourne, Australia; Industrial Transformation Training Centre for Optimal Ageing, Monash University, Clayton, Australia

## Abstract

**Background:** Atrial fibrillation (AF) is associated with aging and increased risk of stroke and dementia. We examined the association between incident AF and cognitive trajectories in older adults using longitudinal data from the ASPirin in Reducing Events in the Elderly (ASPREE) study.

**Methods:** ASPREE was a multi-center randomized, double-blind, placebo-controlled trial of low-dose (100 mg) aspirin in healthy older adults. In this analysis, participants were aged ≥70 years with no history of cardiovascular disease, AF, or a diagnosis of dementia. We used a case ascertainment approach to identify participants with new onset of probable AF over trial participation, defined by new anticoagulant use, incident medically documented diagnosis, or irregular heart rate at study visits prompting diagnosis. We generated *z*-scores for cognitive assessments for global cognition (3MS, Modified Mini-Mental State Examination), verbal fluency/executive function (Controlled Oral Word Association Test, COWAT-F), delayed memory (Hopkins Verbal Learning Test–Revised, HVLT–R), and attention/processing speed (Symbol Digit Modalities Test, SDMT). Baseline cognition and trajectories were compared between probable AF and no AF (control group) using linear mixed-effects models.

**Results:** Of 14,577 participants (mean age = 75.2 ± 4.3 years; 57.8% female), 978 (6.7%) developed probable AF. Baseline cognitive scores did not differ between groups. Compared to controls, those with AF showed greater decline over five years: 0.12 *z*-score (95% CI: 0.05, 0.20) in 3MS, 0.10 (95% CI: 0.07, 0.12) in COWAT, 0.10 (95% CI: 0.10, 0.16) in SDMT, and 0.11 (95% CI: 0.08, 0.14) in HVLT. *APOE ℰ4* allele carriage was associated with greater decline among those with AF.

**Conclusions:** In initially healthy older adults, those who developed AF had comparable cognitive profiles at baseline but exhibited greater cognitive decline over 5 years than those who did not. A diagnosis of AF may be a trigger for cognitive evaluation in older individuals.

**Clinical Trial Registration:** www.clinicaltrials.gov/study/NCT01038583

## Introduction

Atrial fibrillation (AF) is the most commonly sustained cardiac arrhythmia, associated with an increased risk of stroke^1^, heart failure and death^2,3^. The prevalence of AF is highly age-dependent, ranging from 0.1% in those aged under 55 years to 9% in those aged 80 or older^4^, with an expected increase in the USA of at least 2.5-fold by 2050^5^. AF is associated with the development of dementia, independent of shared vascular risk factors, both in those with stroke, including silent cerebral infarcts and cerebral microbleeds^6^, and in the absence of stroke^7^. The predicted increase in AF by 2050 mirrors the predicted 3-fold global increase in dementia from over 57 million cases in 2019 to 153 million cases^8^.

AF was associated with an increased risk of cognitive impairment, with or without a history of clinical stroke in a meta-analysis^9^. Findings from the ARIC study suggested that the association between incident AF with cognitive decline in “stroke-free” individuals may be due to the development of silent stroke, raising the importance of anticoagulation as a strategy to prevent cognitive decline in AF^10^. However, the authors went on to report strong associations between AF and cognitive decline and dementia over 20 years in the ARIC Neurocognitive Study (ARIC-NCS)^11^. Authors of subsequent systematic reviews and meta-analyses have found strong associations between AF and dementia^7,12^, both associated with^6^ and independent of ^13^ incident stroke. There is now encouraging evidence that treatment with direct oral anti-coagulants is associated with a lower incidence of dementia compared to those treated with warfarin, especially with apixaban and rivaroxaban^14,15^. In addition, Mendelian randomization studies have provided novel evidence supporting a potential causal effect of genetically predicted AF on all-cause dementia^16^.

In view of these findings, we aimed to examine associations between cognitive impairment and AF in ASPREE participants. The ASPREE study was a long-term multi-centre double-blind, randomized, placebo-controlled trial investigating the effects of low-dose aspirin in healthy, community-dwelling older adults^1^. Aspirin was not found to have a significant effect on either cognitive trajectory or dementia incidence in the total ASPREE cohort after median 4.7-year follow-up (Ryan, Storey et al 2020). Participants had to be free of an AF diagnosis at study inception. We hypothesized that participants who developed AF would have lower baseline cognition at study entry and have a trajectory of greater cognitive decline than those who do not develop AF. We anticipated that the cognitive domains most affected would include attention, executive function and processing speed, given that these functions are subserved by subcortical processes, most vulnerable to the effects of cerebral small vessel disease (SVD) and hypoperfusion^17-19^.

## Methods

### Study design

The ASPREE study details are available in previous publications^20,21^. In summary, 19,114 adults from Australia and the United States were enrolled between March 2010 and December 2014. Participants were aged ≥70 years (≥65 years for U.S. African American and Hispanic participants) and had no history of cardiovascular disease (including AF), dementia, or a Modified Mini-Mental State examination (3MS) score below 78/100 at baseline, and randomly assigned aspirin (100mg) versus placebo, and followed for a median of 4.7 years. As previously reported, aspirin was not found to have a significant effect on the incidence of dementia, nor on cognitive trajectories^22^. Following the end of the trial, participants were invited to join an observational extension study, ASPREE-XT, wherein assessments mirrored those of the original ASPREE trial. Baseline ASPREE-XT assessments began from February 2018 to October 2019 with annual visits occurring up to April 2024^23^. Ethics approval was obtained from the Monash University Human Research Ethics Committee (2006/745 MC), and all participants provided written informed consent. Sub-study approval for this project was granted in 2019.

### Classification of Atrial Fibrillation (AF) Status

Atrial fibrillation (AF) status was determined using an algorithm applied by research personnel who were blinded to the participants’ treatment allocation and baseline characteristics. This algorithm has been previously described in detail^24^. Overall, participants were classified as having new onset or incident AF within the study if a new diagnosis of AF was identified during the annual review of clinical notes or through self-reports (at 6-month phone call or annual study visit), verified by clinical records. New onset AF was also considered if, on more than one occasion, an irregular heart rate was detected by staff or a self-report of AF was noted along with the prescription of anticoagulants, cardiac glycosides, or other relevant antiarrhythmic medications. All records were classified into three categories: no AF, possible or unconfirmed AF (insufficient evidence of incident AF), and probable AF, where the criteria outlined above were met (referred to as the AF group subsequently; see Supplemental Figure S1 for detail)^25^. AF classification data were collected during the participants’ ASPREE follow-up period (between March 2011 to June 2017).

### Sample Selection

The present study included all ASPREE participants except (1) participants categorized as having possible or unconfirmed AF; and (2) participants aged between 65 and 70 years. Participants aged 65–69 were excluded because this subgroup included only U.S. minorities and no Australian participants, which would limit comparability and consistency of the study population.

### Participant Characteristics and Study Endpoints

Hypertension was through direct BP measures (<140/90) or on treatment for HBP, diabetes was self-report or high FBG or on treatment, high cholesterol was through path report only.

Demographic, social, and lifestyle factors were assessed at trial baseline. These factors included age, years of education, handedness, marital status, family history of stroke or dementia, smoking status, and alcohol consumption (categorized as <1, 1–2, 3–4, 5–6 days per week, and every day). Data on depressive symptoms were obtained through direct assessment, self-reported questionnaires, or prescription medication use at baseline. Hypertension (HTN) was determined by direct blood pressure (BP) measurement (≥140/90 mmHg) or treatment for high blood pressure (HBP). Diabetes mellitus (DM) was defined by self-report, elevated fasting blood glucose (FBG), or treatment for diabetes. Hypercholesterolemia was identified through pathology reports. Body mass index (BMI; kg/m^2^) was calculated using weight and height measurements and classified as underweight (<18.5), healthy weight (18.5–24.9), overweight (25.0–29.9), and obese (≥30). Venous blood samples were collected for APOE genotype determination from participants who provided consent for DNA analysis and storage, and individuals were categorized as either *APOE* ε4 carriers or non-carriers.

Study endpoints included death, stroke, major adverse cardiovascular events (MACE), and dementia. Supporting documents for diagnoses of stroke and MACE were sought from hospitals, specialists and general practice records after incidental reports during the year or at annual follow-up visits; and presented to specialist panels who were masked to randomization and adjudicated cases according to pre-defined criteria^26,27^. Dementia diagnosis based on DSM-IV criteria was determined by clinical specialists using cognitive and functional testing (for those suspected of having dementia), comprehensive clinical assessments, brain imaging and a review of medical history (see Ryan and colleagues^22^ for further details).

### Cognitive Assessment

Cognitive assessments were administered by trained research personnel at baseline and over follow-up (years 0, 1, 3, 5), close-out visits in the final year of the trial (2017) (years 4-7), and at least four additional visits in ASPREE-XT (years 8, 9, 10). The cognitive battery included the 3MS^28^ for global cognitive function, single-letter Controlled Oral Word Association Test (COWAT-F)^29^ for verbal fluency and executive function, Hopkins Verbal Learning Test–Revised (HVLT–R)^30^ for delayed recall, and Symbol Digit Modalities Test (SDMT)^31^, for attention and information processing speed. The scores for each cognitive assessment were converted to standardized *z*-scores, based on a linear predictive model with a spline function (see Supplementary Material for detail). These *z*-scores were adjusted by age at randomization, sex (male vs. female), years of education (<9, 9-11, 12, 13-15, 16, and 17- 21 years), country (Australia vs United States), and self-reported culture (Australian Caucasian vs. U.S. Caucasian vs. U.S. African American vs. U.S. Hispanic vs. Other). The 3MS was developed as an augmented Mini-Mental Status Exam (MMSE), increasing the score range to 100 points, with published normative data for the older individual^32^.

Dementia ascertainment for ASPREE was conducted using additional cognitive testing in participants with suspected dementia via pre-specified triggers, including a 3MS score <78/100, change in score >10.15 points from the 5-year age- and education-adjusted predicted 3MS score, report of cognitive problems to a specialist or clinician diagnosis of dementia, which was conveyed to study staff members, or prescription of cholinesterase inhibitors for presumed AD diagnosis. This was based on within-study assessments or clinical history and adjudicated according to DSM-IV criteria^22^.

### Statistical Analysis

Statistical analyses were performed using R Version 4.3.3. Linear mixed-effects models with random intercepts and slopes were used to assess cognitive trajectories in relation to AF status (no AF vs. AF group) across the follow-up period for each of the four cognitive assessments. For each participant with a diagnosis of stroke, MACE, dementia, or death (i.e., termed an index event), cognitive assessment results following the event, as well as any follow-up assessments where cognitive testing was not conducted, were excluded from the analysis. Missing data, such as missing covariates or cognitive assessment data lost to follow-up, were handled using multiple imputation via the Multiple Imputation by Chained Equations (MICE)^33^. Data were imputed with a wide format using the *missForest* package and reshaped long for analysis.

In these models, the interaction term between follow-up time (defined as 5-year intervals since randomization) and AF status was included to examine whether baseline cognition scores and trajectories differed between participants with no AF and participants in the AF group. Adjusted models were also constructed to account for potential confounders, including hypertension, the criteria of which has been defined previously^34,35^ (yes vs. no), DM (yes vs. no), and BMI categories (underweight, healthy weight, overweight, obese). Sensitivity analyses were conducted by stratifying sex (males and females) and *APOE ε4* status (*ε4* carriers and non-carriers).

Details on all statistical packages, scripts, and functions used are provided in the supplementary material.

## Results

### Participant Characteristics

Table 1 presents the baseline demographic and clinical characteristics and study endpoint data of the sample, categorized by AF status. A total of 14,577 participants were included in the analysis, with a mean age of 75.17 years (SD=4.29) at baseline, and 57.80% were female. Among these participants, 978 (6.71%) were identified as having new-onset AF during the

**Table 1.** Baseline demographic and clinical characteristics, and study endpoint data by AF status.

|  | **No AF**<br>(n=13,599) | **AF Group**<br>(n=978) | **Total**<br>(N=14,577) | *p*-value |
| --- | --- | --- | --- | --- |
| Age at Baseline (years) | 75.1 (4.2) | 76.6 (4.8) | 75.1 (4.3) | <0.001 |
| Duration of follow-up (years) | 7.7 (2.6) | 7.1 (3.1) | 7.7 (2.6) | <0.001 |
| Cultural Background |  |  |  | <0.001 |
| Australian Caucasian | 12006 (88.3) | 897 (91.7) | 12903 (88.6) |  |
| U.S. Caucasian | 751 (5.5) | 53 (5.4) | 804 (5.5) |  |
| U.S, African American | 380 (2.8) | 7 (0.7) | 387 (2.7) |  |
| U.S. Hispanic | 248 (1.8) | 11 (1.1) | 259 (1.8) |  |
| Other | 207 (1.5) | 10 (1.0) | 217 (1.5) |  |
| Sex |  |  |  | <0.001 |
| Male | 5665 (41.7) | 491 (50.2) | 6156 (42.2) |  |
| Female | 7934 (58.3) | 487 (49.8) | 8421 (57.8) |  |
| Education Years |  |  |  | 0.619 |
| <9 Years | 2168 (15.9) | 164 (16.8) | 2332 (16.0) |  |
| 9-11 Years | 4108 (30.2) | 288 (29.4) | 4396 (30.2) |  |
| 12 Years | 1597 (11.7) | 114 (11.7) | 1711 (11.7) |  |
| 13-15 Years | 2216 (16.3) | 175 (17.9) | 2391 (16.4) |  |
| 16 Years | 1253 (9.2) | 91 (9.3) | 1344 (9.2) |  |
| 17-21 Years | 2256 (16.6) | 146 (14.9) | 2402 (16.5) |  |
| Depression Symptoms |  |  |  | 0.049 |
| No | 4045 (75.3) | 167 (68.7) | 4212 (75.0) |  |
| Yes | 1249 (23.2) | 73 (30.0) | 1322 (23.5) |  |
| Unsure | 81 (1.5) | 3 (1.2) | 84 (1.5) |  |
| Hypertension |  |  |  | <0.001 |
| No | 2389 (27.1) | 84 (13.7) | 2473 (26.3) |  |
| Yes | 6370 (72.4) | 527 (85.7) | 6897 (73.2) |  |
| Unsure | 43 (0.5) | 4 (0.7) | 47 (0.5) |  |
| High Cholesterol |  |  |  | 0.033 |
| No | 2883 (42.6) | 128 (35.8) | 3011 (42.2) |  |
| Yes | 3778 (55.8) | 225 (62.8) | 4003 (56.2) |  |
| Unsure | 108 (1.6) | 5 (1.4) | 113 (1.6) |  |
| Diabetes |  |  |  | 0.007 |
| No | 12407 (91.4) | 879 (90.2) | 13286 (91.3) |  |
| Yes | 1129 (8.3) | 87 (8.9) | 1216 (8.4) |  |
| Unsure | 43 (0.3) | 9 (0.9) | 52 (0.4) |  |
| BMI (Classification) |  |  |  | <0.001 |
| Healthy Weight | 3347 (24.7) | 204 (20.9) | 3551 (24.5) |  |
| Underweight | 66 (0.5) | 7 (0.7) | 73 (0.5) |  |
| Overweight | 6252 (46.2) | 421 (43.2) | 6673 (46.0) |  |
| Obese | 3869 (28.6) | 343 (35.2) | 4212 (29.0) |  |
| *APOE ε4* Carrier |  |  |  | 0.780 |
| ε4 non-carrier | 7854 (57.8) | 576 (58.9) | 8430 (57.8) |  |
| ε4 carrier | 2715 (20.0) | 189 (19.3) | 2904 (19.9) |  |
| Not Available | 3030 (22.3) | 213 (21.8) | 3243 (22.2) |  |
| Smoking |  |  |  | <0.001 |
| Current | 479 (3.5) | 27 (2.8) | 506 (3.5) |  |
| Former | 5447 (40.1) | 458 (46.8) | 5905 (40.5) |  |
| Never | 7673 (56.4) | 493 (50.4) | 8166 (56.0) |  |
| Alcohol Consumption |  |  |  | 0.027 |
| < once per week | 3829 (34.0) | 262 (31.7) | 4091 (33.9) |  |
| 1-2 days per week | 1748 (15.5) | 105 (12.7) | 1853 (15.3) |  |
| 3-4 days per week | 1655 (14.7) | 124 (15.0) | 1779 (14.7) |  |
| 5-6 days per week | 1564 (13.9) | 122 (14.8) | 1686 (14.0) |  |
| Every day | 2463 (21.9) | 213 (25.8) | 2676 (22.1) |  |

*Note.* Age at randomization and duration of participation are reported as mean (SD). All other variables are reported as number of participants in the category (%).

follow-up period. Among individuals with no AF, 124 (0.91%) had a stroke, 164 (1.20%) MACE, and 824 (6.06%) dementia. Among individuals in the AF group, 74 (7.57%) had a stroke, 136 (13.91%) MACE, and 71 (7.26%) dementia. The 4,032 participants with possible or unconfirmed AF, and 566 (14.04%) participants aged under 70 years were excluded from this analysis.

### Cognitive Functioning and AF Status

Table 2 shows both unimputed and imputed outcomes from the unadjusted and adjusted linear mixed-effects regression models. There was no evidence of baseline differences in scores on 3MS, COWAT, SDMT, and HVLT-R between participants in the no AF and AF groups. Among all participants over the follow-up period, significant declines were observed in 3MS, SDMT, and HVLT-R scores. These declines were greater among participants with AF. For 3MS, for example, an additional 0.12 *z*-score (95%CI: 0.01, 0.23) deterioration per five years were observed among participants with AF. The largest trend differences were observed in the SDMT, where a further 0.17 *z*-score (95%CI: 0.12, 0.22) deterioration per five years was observed for those with AF compared to those without. While there was a 0.11 *z*-score (95%CI: 0.09, 0.12) increase in the whole sample for the COWAT, the rate of change among participants in the AF group was 0.09 *z*-scores (95%CI: 0.04, 0.14) slower than participants with AF, showing a decreasing trajectory. These findings remained after adjusting for hypertension, presence of DM, and baseline BMI classification. Figure 1 displays the cognitive trajectories by AF status for each cognitive assessment.

**Table 2.** Unimputed and Imputed Results for Unadjusted and Adjusted Linear Mixed Effects Models.

|  |  | Unadjusted Model |  |  |  | Adjusted Model |  |  |  |
| --- | --- | --- | --- | --- | --- | --- | --- | --- | --- |
|  |  | Unimputed |  | Imputed |  | Unimputed |  | Imputed |  |
| Cognitive assessment | Variable | Coefficient (95% CI) | p-value | Coefficient (95% CI) | p-value | Coefficient (95% CI) | p-value | Coefficient (95% CI) | p-value |
| 3MS | Time (5 years) | -1.18<br>(-1.20, -1.16) | <0.001 | -1.44<br>(-1.46, -1.42) | <0.001 | -1.23<br>(-1.26, -1.20) | <0.001 | -1.44<br>(-1.46, -1.42) | <0.001 |
|  | AF Status<br>(reference: No AF) | 0.03<br>(-0.06, 0.13) | 0.461 | 0.07<br>(-0.04, 0.17) | 0.203 | 0.00<br>(-0.12, 0.12) | 0.968 | 0.05<br>(-0.05, 0.16) | 0.349 |
|  | **AF Status & Time Interaction** | **-0.09**<br>**(-0.17, -0.01)** | **0.033** | **-0.12**<br>**(-0.20, -0.05)** | **0.001** | **-0.12**<br>**(-0.23, -0.01)** | **0.028** | **-0.12**<br>**(-0.20, -0.05)** | **0.002** |
| COWAT | Time (5 years) | 0.09<br>(0.09, 0.10) | <0.001 | 0.03<br>(0.03, 0.04) | <0.001 | 0.11<br>(0.09, 0.12) | <0.001 | 0.03<br>(0.03, 0.04) | <0.001 |
|  | AF Status<br>(reference: No AF) | 0.04<br>(-0.02, 0.10) | 0.198 | 0.05<br>(-0.02, 0.12) | 0.161 | 0.05<br>(-0.03, 0.13) | 0.199 | 0.06<br>(-0.01, 0.13) | 0.086 |
|  | **AF Status & Time Interaction** | **-0.08**<br>**(-0.11, -0.04)** | **<0.001** | **-0.10**<br>**(-0.12, -0.07)** | **<0.001** | **-0.09**<br>**(-0.14, -0.04)** | **<0.001** | **-0.10**<br>**(-0.12, -0.07)** | **<0.001** |
| SDMT | Time (5 years) | -0.36<br>(-0.37, -0.35) | <0.001 | -0.48<br>(-0.49, -0.48) | <0.001 | -0.36<br>(-0.37, -0.35) | <0.001 | -0.48<br>(-0.49, -0.48) | <0.001 |
|  | AF Status<br>(reference: No AF) | -0.02<br>(-0.08, 0.05) | 0.619 | -0.05<br>(-0.12, 0.02) | 0.157 | 0.00<br>(-0.08, 0.08) | 0.961 | -0.04<br>(-0.11, 0.03) | 0.257 |
|  | **AF Status & Time Interaction** | **-0.13**<br>**(-0.16, -0.10)** | **<0.001** | **-0.10**<br>**(-0.12, -0.07)** | **<0.001** | **-0.17**<br>**(-0.22, -0.13)** | **<0.001** | **-0.10**<br>**(-0.12, -0.07)** | **<0.001** |
| HVLT-R | Time (5 years) | -0.04<br>(-0.05, -0.03) | <0.001 | -0.15<br>(-0.16, -0.14) | <0.001 | -0.03<br>(-0.04, -0.01) | <0.001 | -0.15<br>(-0.16, -0.14) | <0.001 |
|  | AF Status<br>(reference: No AF) | 0.06<br>(-0.01, 0.12) | 0.086 | 0.07<br>(-0.01, 0.14) | 0.068 | 0.02<br>(-0.07, 0.10) | 0.689 | 0.07<br>(-0.01, 0.14) | 0.072 |
|  | **AF Status & Time Interaction** | **-0.07**<br>**(-0.11, -0.04)** | **<0.001** | **-0.11**<br>**(-0.14, -0.08)** | **<0.001** | **-0.09**<br>**(-0.14, -0.05)** | **<0.001** | **-0.11**<br>**(-0.14, -0.08)** | **<0.001** |

*Note.* 3MS = Modified Mini-Mental State examination, COWAT = Controlled Oral Word Association Test, HVLT-R = Hopkins Verbal Learning Test Revised, SDMT = Symbol Digit Modalities Test, AF = Atrial Fibrillation. The models were adjusted by hypertension (yes vs. no), diabetes mellitus (DM; yes vs. no), and body mass index (BMI) categories (underweight, healthy weight, overweight, obese). Outcome z-scores were standardised by age at randomization, sex, years of education), country, and culture.

**Figure 1.**
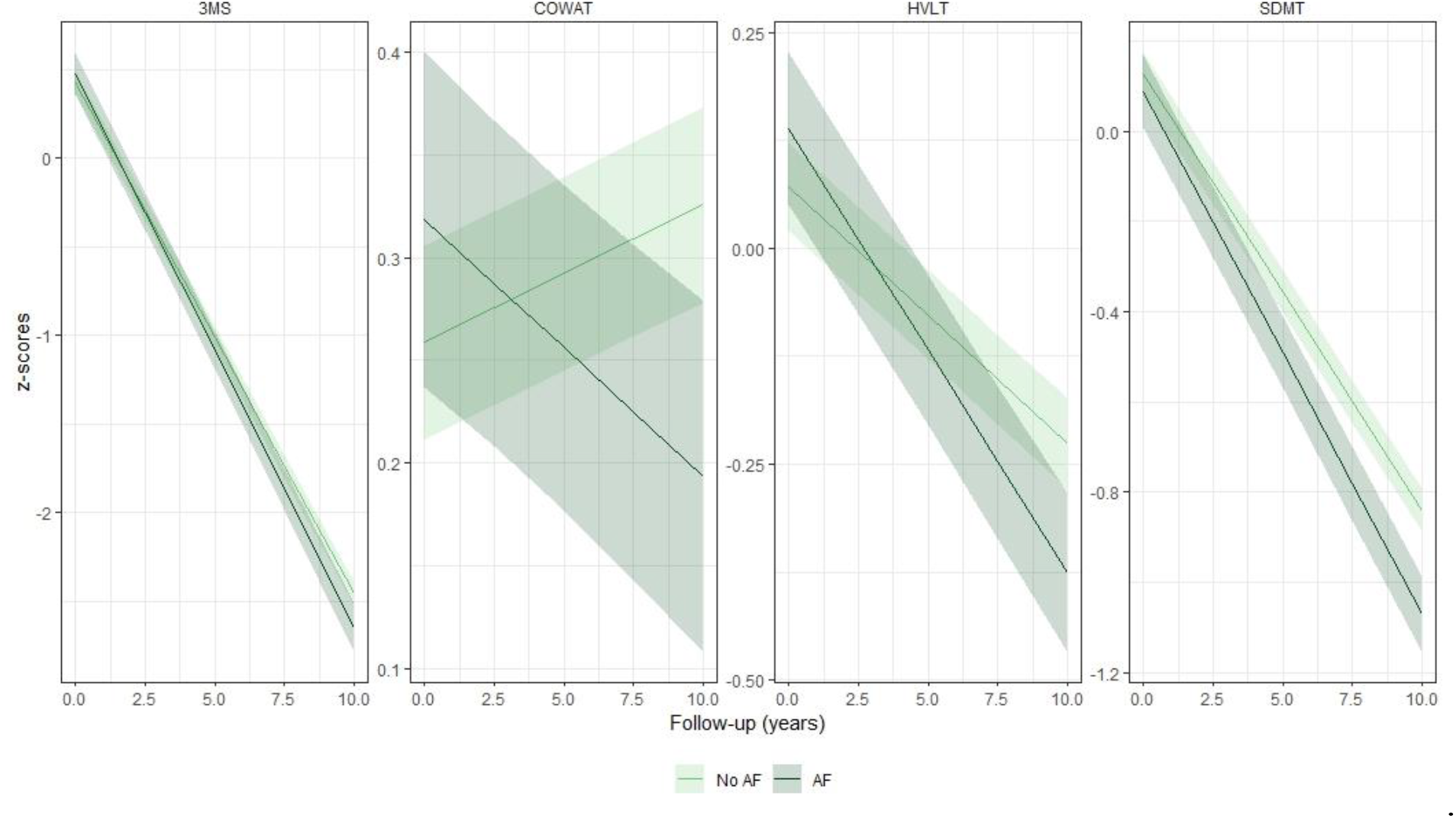
Marginal means plots displaying the predicted trajectories of global cognition (Modified Mini-Mental State Examination; 3MS), verbal fluency and executive functioning (Controlled Oral Word Association Test; COWAT), delayed recall (Hopkins Verbal Learning Test; HVLT-R), and attention and information processing speed (Symbol Digit Modalities Test; SDMT) over a 10-year period for individuals who developed incident AF during the ASPREE follow-up period and those who did not, based on the adjusted imputed results from Table 2.

### Sensitivity Analyses by Participant Sex

Supplementary Table S1 presents the unadjusted and adjusted linear mixed effects models for female participants only, while Supplementary Table S2 shows the results for male participants only. Figure 2(A) illustrates the coefficients for the interaction between time and AF status for males and females. The trend observed in the total population remained comparable between male and female participants, except for the 3MS, where the difference in cognitive trajectories between female participants with and without AF was not statistically significant.

**Figure 2.**
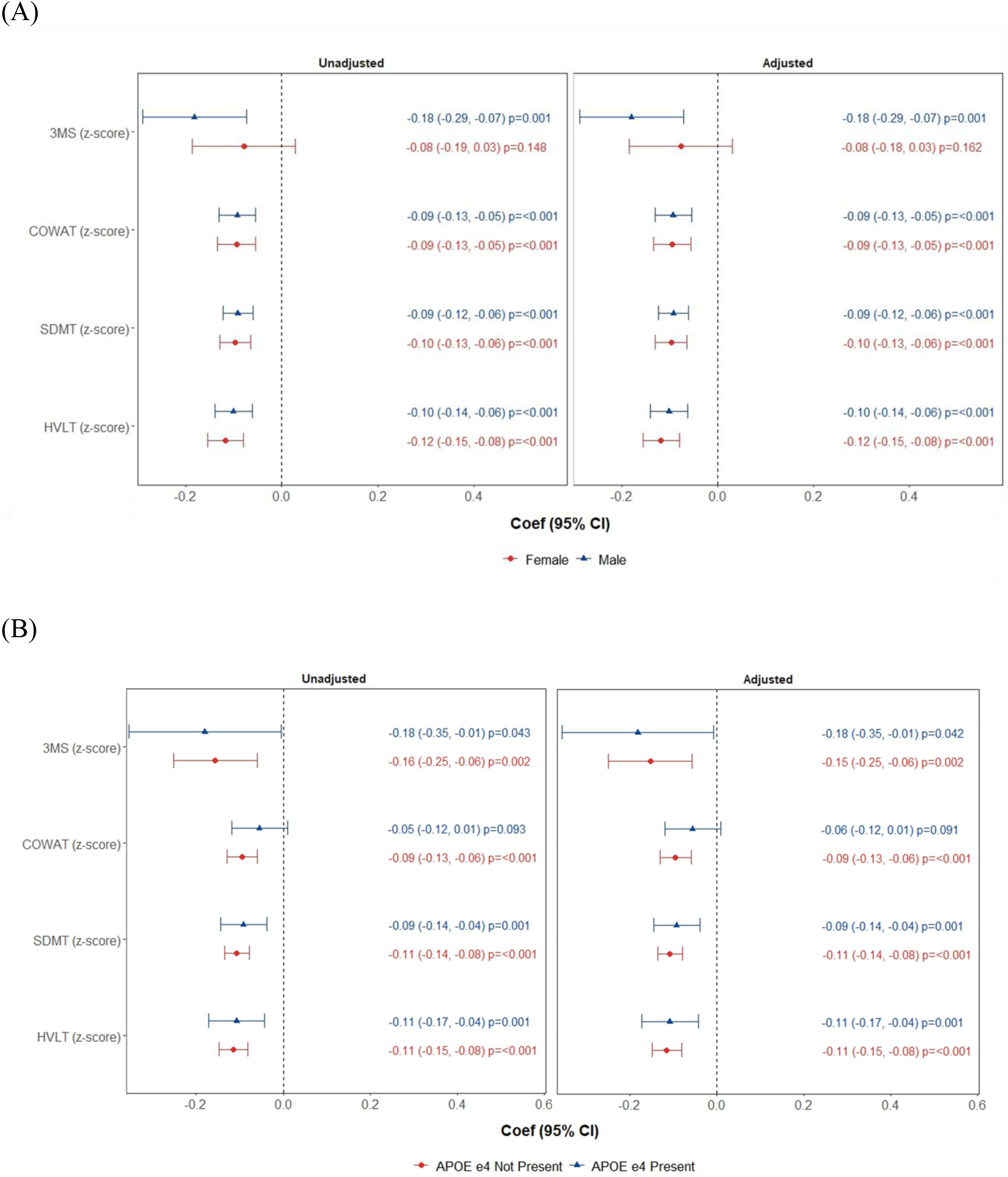
Forest plot displaying the coefficients for the Time by AF Status Interaction by (A) Sex, and (B) *APOE ε4* status using imputed data. *Note*. 3MS = Modified Mini-Mental State examination, COWAT = Controlled Oral Word Association Test, HVLT-R = Hopkins Verbal Learning Test Revised, SDMT = Symbol Digit Modalities Test. Outcome *z*-scores were standardized by age at randomization, sex, years of education, country, and culture. The interaction coefficient represents the differential rate of cognitive change over five-year increments between individuals with AF and those without. Specifically, it quantifies whether the presence of AF is associated with a faster or slower rate of cognitive decline compared to those who do not develop AF. In this case, individuals with AF exhibited a faster rate in cognitive decline in all cognitive measures, regardless of sex and *APOE ε4* carriage status, with the potential exception being 3MS (global function) amongst females.

### Sensitivity Analyses by APOE ε4 Carriers and Non-Carriers

Supplementary Table S3 presents the unadjusted and adjusted linear mixed-effects for *APOE ε4* carriers, while Supplementary Table S4 shows the results after *APOE ε4* non-carriers. Figure 2(B) illustrates the coefficients for the interaction between time and AF status for *APOE ε4* carriers and non-carriers. Overall, the greater rate of decline in cognitive trajectories amongst those with AF was similar amongst *APOE ε4* carriers and non-carriers.

## Discussion

This study explored the longitudinal relationship between new-onset atrial fibrillation (AF) and cognitive trajectories in healthy older adults using data from a large longitudinal follow-up study. Our findings suggest that while cognitive performance at baseline was similar between individuals who did or did not develop AF, those who developed AF experienced a greater decline across all assessed cognitive domains over time, including global cognition (3MS), executive function (COWAT), attention and processing speed (SDMT), and memory recall (HVLT-R). The results underscore the clinical importance of monitoring cognitive function in older adults diagnosed with AF, suggesting that new-onset AF is a potential marker for accelerated cognitive decline. These findings align with previous research associating AF with cognitive impairment and reinforce the need for proactive cognitive evaluations and targeted interventions for individuals with AF, including the importance of anticoagulation.

Participants who developed AF during the ASPREE study were older, more likely to be male and Caucasian, and to have a higher BMI. Unsurprisingly, vascular risk factors, including hypertension, hypercholesterolemia, history of smoking and diabetes mellitus, were associated with incident AF. Daily alcohol, but not less frequent, consumption was associated with development of AF. Those who developed AF were more likely to have incident stroke or MACE. Proportionately more participants in the AF group were assigned a dementia diagnosis than those without AF (7.3% versus 6.1%) although the difference was not statistically significant.

Amongst the individual measures of cognitive performance, the COWAT scores showed a slight increase overall, explicable via practice or learning effects in this cohort, as the same letter was used for each annual assessment^36^. Approaches to counter this have been proposed in longitudinal studies of preclinical AD^37,38^. Interestingly, this increase was not observed in those with AF, suggesting a flattening or attenuation of learning which has been previously described in people with preclinical AD or MCI^39,40^.

Carriage of the *APOE ε4* allele has been shown to affect cortical thinning in normal aging^41^ and in cerebrovascular disease^42^. In our study, greater cognitive decline associated with AF was observed within the *APOE ε4* carrier group. This pattern is consistent with prior reports suggesting that genetic risk and vascular factors may have additive effects on cognitive impairment and cognitive decline in *APOE ε4* carriers with vascular risk factors^43^. Recent imaging studies have highlighted this interaction: Petersen et al. found attention, executive, reasoning dysfunction and reduced processing speed in 1335 people with isolated AF without a previous stroke^44^. Importantly, these cognitive deficits were correlated with thinner cortices and decreased metrics of white matter integrity. We have previously reported an association between AF, brain volume and cognitive impairment after stroke^45^, as well as the effects of vascular risk factors and disease including AF on executive function and white matter microstructural changes^19^ in a cohort of ischemic stroke survivors.

The absence of a baseline difference between the AF and no-AF groups strengthens our findings of different cognitive trajectories in our participant groups. There was a 1.44 decrease in *z*-score in our global cognitive measure, the 3MS, for every five years of follow-up for no-AF ASPREE participants, with an additional 0.12 *z*-score decrease per five years among participants with AF. The largest trend differences were observed in our measure of attention and processing speed, the SDMT, in which a 0.13 *z*-score decrease per five years was observed in the AF group compared with the non-AF group. This is consistent with an hypothesis that cognitive functions dependent on white matter health (e.g., reaction time, processing speed) and distributed brain networks (e.g., attention, executive function) would be most associated with the development of AF ^46^.

In the ARIC Neurocognitive Study, there was an average greater decline in the global cognitive *z*-score of -0.115 (95% confidence interval, 0.014–0.215) in participants with AF than in those without AF over 20 years ^11^; i.e., comparable to the z-score=-0.12 we report. They also found an association with dementia (2106 participants developed AF and 1157 participants developed dementia), perhaps because of their longer follow-up. Participants in the ARIC-NCS were younger at inception (56.9 years, 56% women, 24% black) and an AF diagnosis was not an exclusion criterion. Indeed, their cohort may have been enriched for cardiovascular disease risk factors. Their cognitive tests examined similar domains: Delayed Word Recall Test for verbal learning and episodic memory at a 5-minute delay, Digit Symbol Substitution Test for executive function and processing speed, and Word Fluency Test for executive function and expressive language. Ascertainment of dementia was also by a more formalized process, including the administration of MMSE, CDR-SOB and a functional questionnaire, which may have captured more people with dementia than ASPREE dementia ascertainment processes^22^.

The decline in the verbal memory measure, the HVLT-R, is notable. Verbal episodic memory impairments are a pathognomonic feature of Alzheimer disease (rapid forgetting with axial memory deficits) but can be seen in vascular cognitive impairment and dementia, albeit with retrieval deficits. As the delayed and recognition memory scores were combined in the ASPREE cohort, memory impairments could be associated with either cause. AF has been linked to memory decline in stroke survivors^47^ but episodic memory impairments have been described in stroke-free people with AF^48^. In the ARIC-NCS, AF was associated with significantly greater decline in the scores of all tests except the DWRT, their test of verbal episodic memory or delayed recall^11^. The differences noted here could be due to the older age of our participants (median 75.2 years at study inception) and because AF was an enrolment exclusion criterion.

Strengths of our study include the protocol-driven phenotyping of the cohort, in person delivery of an extensive cognitive battery by accredited research staff, greater representation of female participants, exclusion of AF at study inception to allow observations in those who go on to develop AF, long duration of follow-up, and older age at study entry. Our novel and meticulous statistical approach is also a study strength, allowing us to disentangle the potential effects of AF developing during study participation.

However, there are several limitations. Results from the ASPREE cohort are predominantly generalizable to a White/Caucasian population consisting mostly of more educated individuals from high socioeconomic locations. It is possible that more subtle cognitive deficits were not detected by the tools used in ASPREE. There are also potential effects of recruitment bias, as participants were pre-screened for the presence of cognitive impairment. AF was ascertained using a probabilistic algorithm, meaning some people with AF could have been excluded from our probable AF group, and vice versa. We cannot be certain that participants did not have subclinical or asymptomatic AF that only became clinically evident following their baseline visit. Administration of the cognitive tests was done after extensive training of study staff, but included the same tests administered at each visit without alternative versions. Dementia triggers relied on these tests plus clinical triggers over trial participation, and a unique battery of additional tests was performed during a dedicated ‘dementia assessment’ visit, and the participant’s primary care provider notified of the outcomes. Nonetheless, ASPREE did not refer for formal assessment and syndromic clarification was not performed. Participants who may have developed dementia but missed their assessments, or whose clinical history did not include these triggers, would not have been captured. In addition, although incident clinical stroke was imputed, systematic brain imaging was not done routinely, meaning that the effects of silent brain infarction or white matter hyperintensities were not estimated.

## Conclusions

In a cohort of initially AF-free, cognitively normal older participants at study entry, we found that those who developed AF had greater cognitive decline than those who did not, despite entering the study with comparable cognitive profiles. Participants who developed AF were older, with more vascular risk factors and were more likely to be male and Caucasian. Cognitive domains affected were processing speed, attention, executive function and memory. Among *APOE ℰ4* carriers, those with AF showed greater cognitive decline than those without AF, consistent with an additive effect of genetic and vascular risk. A diagnosis of AF should form a trigger for cognitive evaluation and monitoring in older individuals.

## Data Availability

The data that support the findings of this study are available from the corresponding author upon reasonable request.

## Acknowledgements

The authors gratefully acknowledge the dedicated and skilled staff in Australia and the United States whose contributions made the ASPREE clinical trial possible. We are especially grateful to the ASPREE participants for their generous involvement, and to the medical staff and clinics who provided their care.

